# MAGNITUDE OF NEONATAL HYPOGLYCEMIA AND ITS ASSOCIATED FACTORS AMONG NEONATES ADMITTED TO NEONATAL INTENSIVE CARE UNIT AT HAWASSA CITY PUBLIC HOSPITALS, ETHIOPIA, 2023

**DOI:** 10.1101/2024.07.01.24309773

**Authors:** Selam Tadele, Wegene Jembere, Mastewal Aschale, Tewodros Mulugeta, Samuel Jigso, Mequanint Ayehu, Migbar Sibhat

## Abstract

**Background:** Neonatal hypoglycemia is one of the most common metabolic abnormalities seen in newborns. It is a major contributing factor to neonatal morbidity and mortality. Globally, it affects around 5–15% of all babies and approximately 50% of at-risk babies.. In Ethiopia, neonatal hypoglycemia is frequently diagnosed and one of the commonest causes of admission to the neonatal intensive care unit. Nevertheless, documented records regarding its magnitude and factors associated with hypoglycemia are scarce in the study area. Therefore we aimed to assess the magnitude of neonatal hypoglycemia and its associated factors among neonates admitted to the neonatal intensive care unit at Hawassa City Public Hospitals, Ethiopia.

**Method:** Institution-based cross-sectional study was conducted from April 20 – June 20, 2023among 293 neonates. A systematic random sampling technique was used to reach the study subjects. The data was collected through face-to-face interviews and card review by using structured pretested questionnaire and analyzed using SPSS software version 25. A multivariable logistic regression model was used to determine factors significantly associated with neonatal hypoglycemia with adjusted odds ratio, p-values <0.05 at 95% confidence interval (CI).

**Result:** The magnitude of neonatal hypoglycemia was found 16.6%. Variables significantly associated with the occurrence of neonatal hypoglycemia were: Diabetes mellitus [AOR=9.8, 95%CI (3.08-31.37)], perinatal asphyxia [AOR=2.87, 95%CI (1.07-7.72)], delayed initiation of breastfeeding [AOR=2.63, 95%CI (1.04-6.6)] and hypothermia [AOR=3.8, 95%CI (1.6-9.1)].

**Conclusion:** In this study the magnitude of neonatal hypoglycemia among neonates was high. Neonates with hypothermia, perinatal asphyxia, and delayed initiation of breastfeeding and maternal history of diabetes mellitus have an increased risk of developing hypoglycemia. Hence, Health care providers who are working on delivery and neonatal care should focus on early identification and management of these identified factors.

## 1. Introduction

Neonatal hypoglycemia is defined as an abnormally low blood glucose level (1). There has been much controversy about the ‘numerical’ definition of neonatal hypoglycemia. Although the most often used definition of newborn hypoglycemia is a glucose concentration of 47 mg/dl (2.6 mmol/l), (2, 3), in Ethiopia it is clinically defined as a blood glucose level less than 40 mg/dl (4)..

It is the most prevalent metabolic abnormality in the newborn period (5), which is a major contributing factor to neonatal morbidity and mortality(6). Globally, it affects around 5–15% of all babies (7, 8) and approximately 50% of at-risk babies and is associated with a range of adverse consequences (9)..

In resource-limited settings, evidence shows there is widespread neonatal hypoglycemia, even though in some countries there is a shortage of advanced techniques for hypoglycemia detection and an absence of routine assessment for hypoglycemia. In African countries Nigeria and Uganda have an incidence of 32.7% and 2.2 % respectively (6, 10). In Ethiopia, the magnitude ranges from 15%-25% (11, 12).

If left untreated, severe and prolonged hypoglycemia can have a devastating neurologic and developmental outcome which includes cerebral palsy, long-term neurodevelopmental disabilities, mental retardation, epilepsy, personality disorders, and even death (13).

Despite these devastating health impacts of neonatal hypoglycemia, organized data regarding the magnitude of hypoglycemia and the factors that predict its occurrence were scarce in Ethiopia. Furthermore, we couldn’t find a single study to investigate this issue in our study area. Hence, this study aims to determine the magnitude of neonatal hypoglycemia, and factors associated with it among neonates admitted to neonatal intensive care units at Hawassa City Public Hospitals, Sidama Regional State, Ethiopia.

## 3. Methods and Materials

### 3.1. Study design and area

An institutional-based cross-sectional study was conducted in Hawassa City from April 20-June 20, 2023. Hawassa city is the capital city of the newly emerged Sidama National Regional State and it is located 275 km south of far from Addis Ababa, Ethiopia. There are 9 hospitals (4 public and 5 private) in Hawassa City, as well as 12 public health centers and 18 health posts. The four public hospitals are Hawassa University Comprehensive Specialized Hospital (14), Adare General Hospital (15), Motite Furra Primary Hospital and Hawela Tula Primary Hospital (HTPH). Only the three hospitals have NICU. All three hospitals follow Ethiopia’s National Standard Treatment Guideline for Hospitals(16).

### 3.2. Source population and Study population

#### Source population

- All neonates who were admitted to the neonatal intensive care units of Hawassa city public hospitals are the source population

#### Study population

- All randomly selected neonates admitted to the neonatal intensive care unit of selected public hospitals during the study period were the study population

### 3.3. Inclusion and Exclusion Criteria

All neonates who were admitted to NICU in the selected hospitals during the data collection period were included and neonates admitted without mothers or caregivers were excluded from the study.

### 3.4. Sample size calculation

The sample size was determined by using single population proportion formula by taking the prevalence of neonatal hypoglycemia from the previous study done in Hiwot Fana Hospital, East Harerge, Ethiopia which is 21.2% (11). Considering the parameters of the single population proportion formula which are a marginal error of 0.05, a 95% confidence interval, and a p-value of 0.21 assuming a 15% non-response rate.

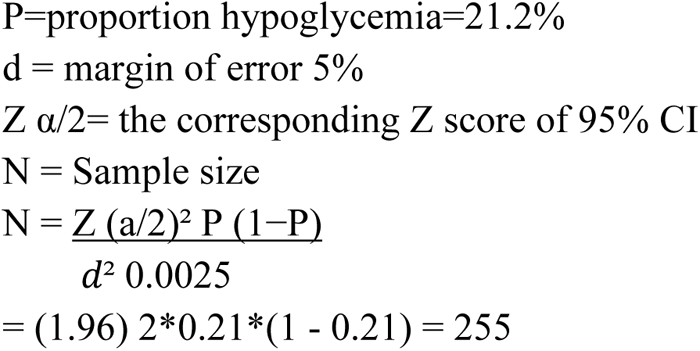

By adding a 15 % non-response rate, a total of 293 participants were included in the study.

### 3.5. Sampling technique

Three public hospitals (Hawassa University Comprehensive Specialized Hospital, Adare General Hospital, and Tulla Primary Hospital) are found in Hawassa City which had NICU. Motite Furra Primary Hospital was excluded because it does not provide NICU service. To select the study participants from each hospital, the proportional allocation formula was used based on caseloads by fixing two months of admission before the survey. Simple random sampling was used to select the first participant and systematic random sampling was used for the next consecutive samples.

### 3.6. Operational definition

**Magnitude of hypoglycemia**: The number of neonates who develop hypoglycemia at admission to NICU during the study period.

**Neonatal hypoglycemia:** - A measure of blood glucose level less than 40 mg/dl(4).

**ANC follow up**: any history of visit or follow up during current pregnancy at any health institution for check-up of pregnancy recorded on the chart.

**Hypothermia:** A neonate’s axillary body temperature below 36.5 o C(4).

**Perinatal asphyxia:** was considered when the 5th minute APGAR score is less than 7 or a neonate did not cry immediately at birth or resuscitation needed at birth and documented on the chart(4).

### 3.7. Data collection tools and procedures

The data was collected using the tool adopted from the previous studies done at Addis Abeba (Tikur Anbessa and St., Paulos), and Harar, Hiwot Fana Hospital (Fikirte Kasaye, 2021). A structured interviewer-based questionnaire and structured checklist was used to collect data for maternal socio-demographic factors and obstetric-related characteristics, and neonatal related factors was taken using a structured checklist from the medical records of a neonate during the data collection period. The data was collected from 20^th^ April, 2023 to June 20, 2023.

Regarding the blood glucose level of the neonate, a blood sample was taken from the neonate, and their blood glucose level was measured using a glucometer. Blood glucose was determined using the Precisa Active Blood Glucose Meter at the admission. Blood glucose results were provided in mg/dl. Neonates who were found to be hypoglycemic were managed as per the NICU protocol. For the data collection, three data collectors and one supervisor who were BSC nurses and who had research experience were chosen. Before data collection one-day training was given for data collectors and supervisors.

### 3.8. Data management and analysis

Data was checked for completeness and consistency and then it was cleaned, coded and entered using Epi data version 3.1 and it was exported to SPSS software version 25 for analysis. Cross-tabulation was done among dependent variables and independent variables. Frequencies, Percentage, mean and standard deviation were used to summarize descriptive statistics. Binary logistic regression and multiple logistic regression were done at p-value <0.25 and <0.05 respectively with a 95% Confidence interval. Hosmer and Lomenshow goodness of fit test was done to check model fitness and the model was fitted. In addition, a multi-collinearity or tolerance test was done with all variables to solve the issue of confounding effects among independent variables. Finally tables and charts were used for data presentation.

### 3.9. Quality assurance technique

The questionnaire was prepared in English and translated to Amharic and sidammu-afu and then retranslated to English to check for inconsistency. The data collection tool was pretested on 5% two weeks before the actual data collection period and modifications were made accordingly. The trained data collectors were assigned to selected hospitals and they collected the data.

During the data collection time, close supervision and monitoring was carried out by supervisors and investigator to ensure the quality of the data. Daily evaluations of the data for completeness and encountered difficulties at the time of data collection were attended accordingly. Finally, all the collected data was checked by the supervisor and investigator for completeness and consistency during the data management, storage, and analysis.

### 3.10. Ethical considerations

Ethical clearance was obtained from the Hawassa University College of Medicine and Health Science institutional review board (Ref Number: IRB/313/15). Then officials at different levels in the Hawassa city administration health office and hospitals were communicated through letters. Since neonates could not speak any information, the neonates’ mothers or guardian were informed about the purpose of the study and written informed consent was obtained to confirm willingness. They were notified that they had the right to refuse or terminate at any point of the interview. The study was conducted as per the declaration of Helsinki. Confidentiality of the information was secured throughout the study process.

## 4. Result

### 4.1. Descriptive statistics of the result

#### 4.1.1. Socio-demographic characteristics of the respondents

A total of 271 neonates were included with an overall response rate of 92.5%. The mean age of the mothers was 29.1 (±5.29) years. The majority of participants were living in urban areas 179(66.6%). Regarding the educational status of the mothers, a higher proportion of mothers 134(49.5%) completed secondary educational level. Concerning the marital status, 244 (90%) mothers were married. Whereas, five (1.8%) and thirteen (4.8%) mothers were widowed and divorced respectively. (Table 1)

**Table 1:**
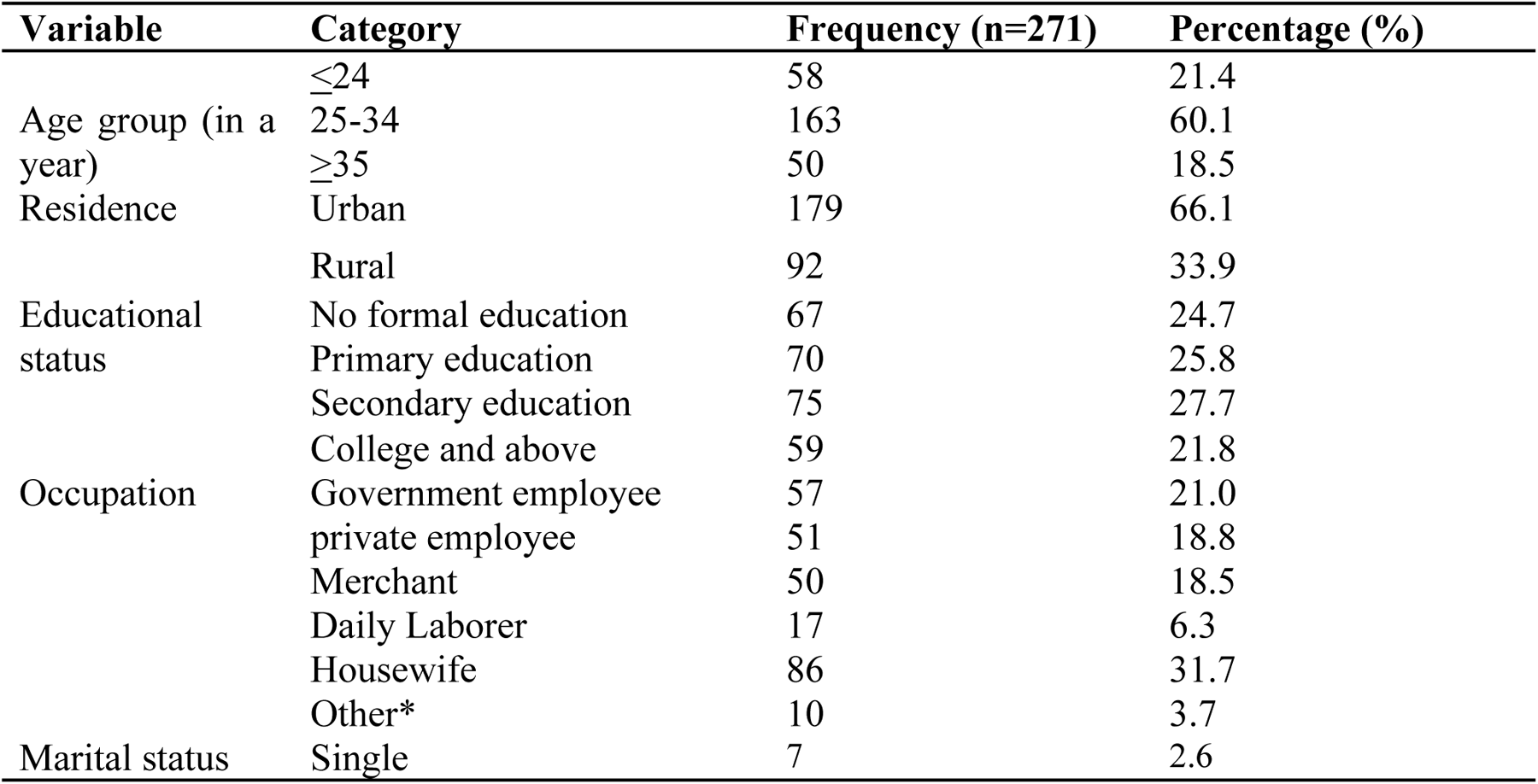
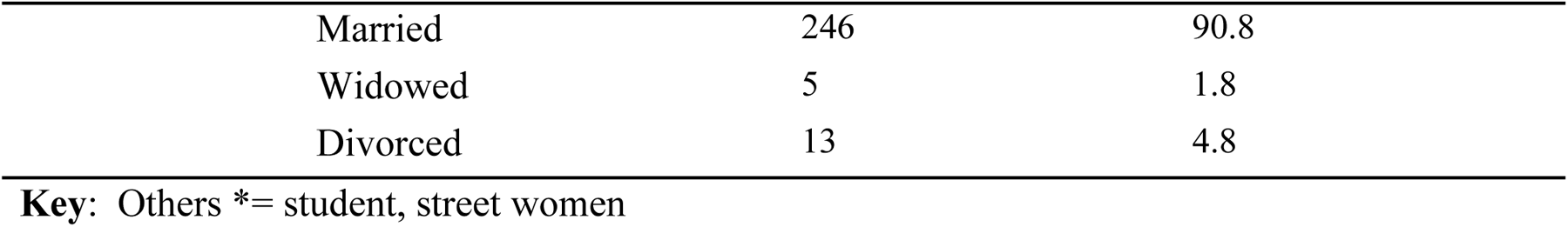
Socio-demographic characteristics of the mothers of neonates on admission to the neonatal intensive care unit in Hawassa City Public Hospitals, Ethiopia 2023. (n=271)

| Variable | Category | Frequency (n=271) | Percentage (%) |
| --- | --- | --- | --- |
| Age group (in a year) | $\leq 24$ | 58 | 21.4 |
|  | 25-34 | 163 | 60.1 |
| | $\geq 35$ | 50 | 18.5 |
| Residence | Urban | 179 | 66.1 |
|  | Rural | 92 | 33.9 |
| Educational status | No formal education | 67 | 24.7 |
|  | Primary education | 70 | 25.8 |
|  | Secondary education | 75 | 27.7 |
|  | College and above | 59 | 21.8 |
| Occupation | Government employee | 57 | 21.0 |
|  | private employee | 51 | 18.8 |
|  | Merchant | 50 | 18.5 |
|  | Daily Laborer | 17 | 6.3 |
|  | Housewife | 86 | 31.7 |
|  | Other* | 10 | 3.7 |
| Marital status | Single | 7 | 2.6 |

|  |  |  |
| --- | --- | --- |
| Married | 246 | 90.8 |
| Widowed | 5 | 1.8 |
| Divorced | 13 | 4.8 |

**Key:** Others \*= student, street women

#### 4.1.2. Description of maternal and obstetric characteristics

In this study, the majority of 234(86.3%) of the mothers had ANC follow-up during the current pregnancy. Of these, 115(45.8%) had at least four ANC follow up. About 194 (71.9%) mothers had more than one child. Regarding the place of delivery, the majority of the neonates were delivered at health institutions. of which 221(72.3%) were delivered at the hospital followed by 59(21.8%) at health centers. Concerning obstetrics history, 14(5.2%) of the mother had prolonged labor and higher proportions of the neonate 166(61.6%) were delivered through spontaneous vaginal delivery. Moreover, 121(44.6%) and 122(45%) of the delivery process were conducted by midwives and medical Doctors (general practitioners, residents and specialists) respectively (Table 2).

As shown in Table 2, Around 26(9.6%) of mothers had diabetes mellitus. Among those 13 (4.8%) of the mothers had gestational Diabetes mellitus and 13 (4.8%) of them had pre-gestational diabetes mellitus. Regarding pregnancy-induced hypertension, 41(15.1%) of mothers had developed it and 20(7.4%) of the mothers were preeclamptic. Regarding other chronic illnesses, 15(5.5%) of the mothers were hypertensive and 10(3.7%) were HIV/AIDs clients (Table 2).

**Table 2:** Obstetric-related factors of neonatal hypoglycemia among neonates on admission to neonatal intensive care unit in Hawassa City public Hospitals, Ethiopia, 2023. (n=271)

| Variable | Category | Frequency (n=271) | Percentage |
| --- | --- | --- | --- |
| ANC | Had no ANC follow-up | 37 | 13.7 |
|  | Had 1-3 ANC follow-up | 115 | 42.4 |
|  | Had >4 ANC follow-up | 119 | 43.9 |
| Number of children(parity) | 1 child(Primi-parity) | 77 | 28.4 |
|  | More than 1 children (Multi-parity) | 194 | 71.9 |
| Prolonged labor | Yes | 14 | 5.2 |
|  | No | 257 | 94.8 |
| Place of delivery | Home | 16 | 5.9 |
|  | Health Center | 59 | 21.8 |
|  | Hospital | 196 | 72.3 |
| Position | Cephalic | 221 | 81.5 |
|  | Non-cephalic | 50 | 18.5 |
| Mode of delivery | Spontaneous vaginal delivery | 166 | 61.3 |
|  | Assisted vaginal delivery | 31 | 11.4 |
|  | Cesarean Section | 74 | 27.3 |
| Premature rupture of membrane | Yes | 58 | 21.4 |
|  | No | 213 | 78.6 |
| Type of pregnancy | Single | 251 | 92.6 |
|  | Twin and above | 20 | 7.4 |
| Conducted by | Nurse | 12 | 4.4 |
|  | Midwives | 121 | 44.6 |
|  | Medical Doctor | 122 | 45 |
|  | Traditional birth attendant | 16 | 5.9 |
| Diabetes Mellitus | Yes | 26 | 9.6 |
|  | No | 245 | 90.4 |
| Pre-pregnancy DM | Yes | 13 | 4.8 |
|  | No | 258 | 95.2 |
| Gestational DM | Yes | 13 | 4.8 |
|  | No | 258 | 95.2 |
| Pregnancy-induced hypertension | Yes | 41 | 15.1 |
|  | No | 230 | 84.9 |
| Preeclampsia | Yes | 20 | 7.4 |
|  | No | 251 | 92.6 |
| Eclampsia | Yes | 31 | 11.4 |
|  | No | 240 | 88.6 |
| Chronic Hypertension | Yes | 15 | 5.5 |
|  | No | 256 | 94.5 |
| HIV/AIDs | Yes | 10 | 3.7 |
|  | No | 261 | 96.3 |

#### 4.1.3. Neonatal-related variables

Among admitted neonates around 156 (57.6%) were male and more than half 171 (63.1%) were gestational age between 37-42 weeks. About 112 (41.3%) of them had birth weights between 2500gm-4000gm. The majority of the neonates 216 (79.7%) were admitted within 24hr after delivery. Around, 119 (43.9%) of the neonates had hypothermia. Regarding feeding initiation time 99 (36.5%) of neonates started feeding within one hour (Table 3).

Concerning the neonatal-related comorbid illness, 241(88.9%) of neonates were admitted with comorbid illness. Among those 130(48%) had neonatal sepsis, 52(19.2%) had birth asphyxia, 38(14%) had respiratory distress syndrome, 30(11.1%) had Meconium Aspiration Syndrome and 53(19.6%) had jaundice.

**Table 3.**
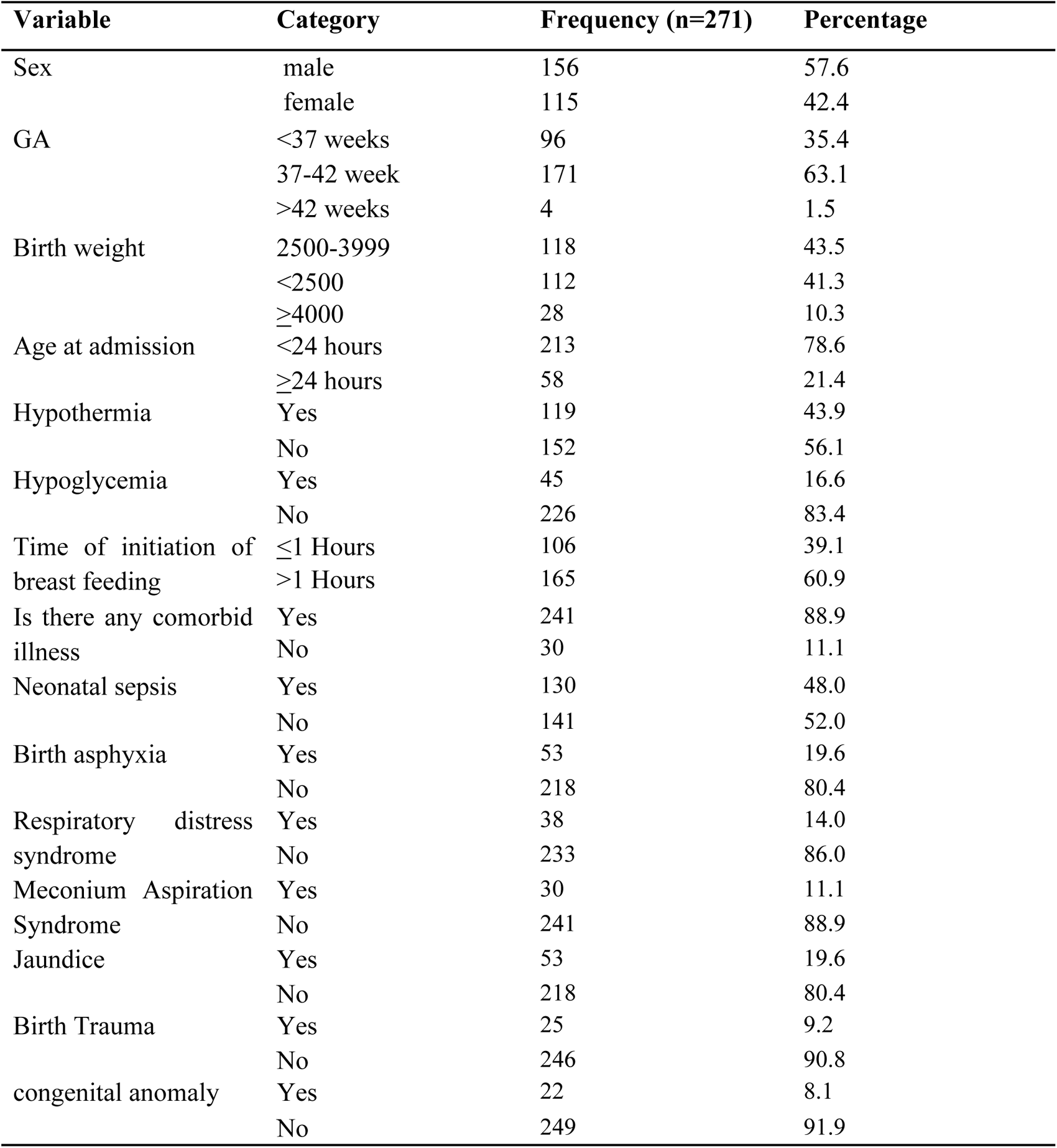
Neonatal-related predictors of hypoglycemia among neonates on admission to neonatal intensive care unit in Hawassa City public Hospitals, Ethiopia 2023. (n=271)

### 4.2. The Magnitude of neonatal hypoglycemia

In this study, the overall magnitude of neonatal hypoglycemia was observed to be 45(16.6%) (**Figure 1**).

**Figure 1.**
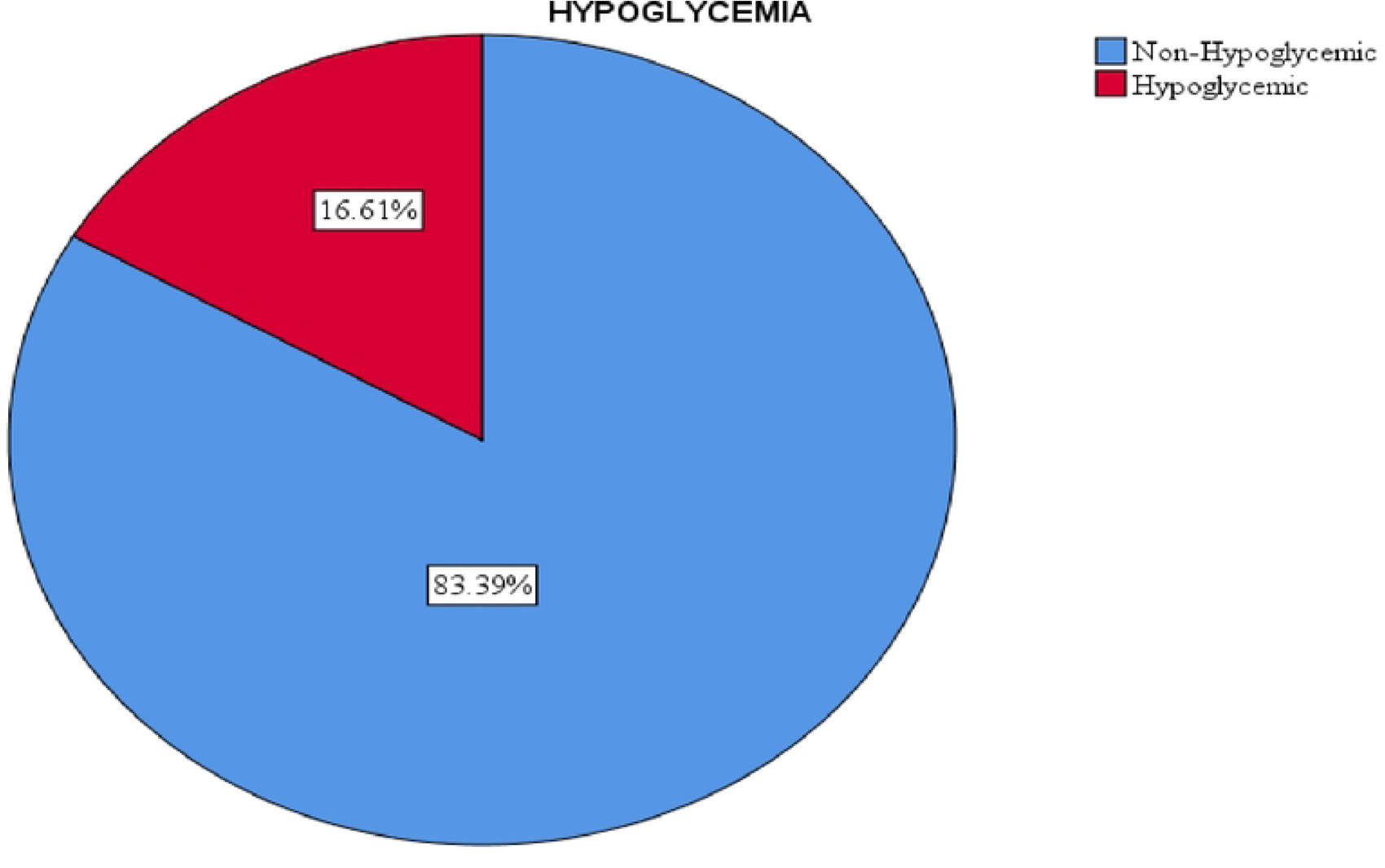
The magnitude of hypoglycemia among neonates on admission to neonatal intensive care uni in Hawassa City Public Hospitals, Ethiopia 2023. (n=27l)

### 4.3. Factors associated with Neonatal Hypoglycemia

The bivariate logistic regression showed that diabetes mellitus, parity, antenatal care visits, prolonged labor, and preeclampsia were selected among maternal-related factors. In the same way, the age of the neonate, birth weight, neonatal sepsis, birth asphyxia, hypothermia, and time of initiation of breastfeeding were selected from neonatal-related risk factors.

The multivariable logistic regression result showed that those mothers who have had Diabetes mellitus were about ten times to have newborns suffering from neonatal hypoglycemia compared with those who had not [AOR=9.8, 95% CI(3.08-31.37)]. Neonates born with Birth asphyxia were nearly three times more likely to be hypoglycemic than their counterpart [AOR=2.87, 95% CI (1.07-7.72)]. Regarding the time of initiation of breastfeeding, neonates who start breastfeeding after one hour of delivery were approximately three times more likely to develop neonatal hypoglycemia than those who start within one hour [AOR=2.63, 95% CI(1.04-6.6)]. Similarly, neonates who develop neonatal hypothermia were nearly four times to be hypoglycemic than their counterparts [AOR=3.8, 95%CI (1.6-9.1)]. (Table 4)

Antenatal care visits, parity, prolonged labor, preeclampsia, birth weight, neonatal sepsis, and age at admission were not found to have statistically significant association with neonatal hypoglycemia in this study.

**Table 4.**
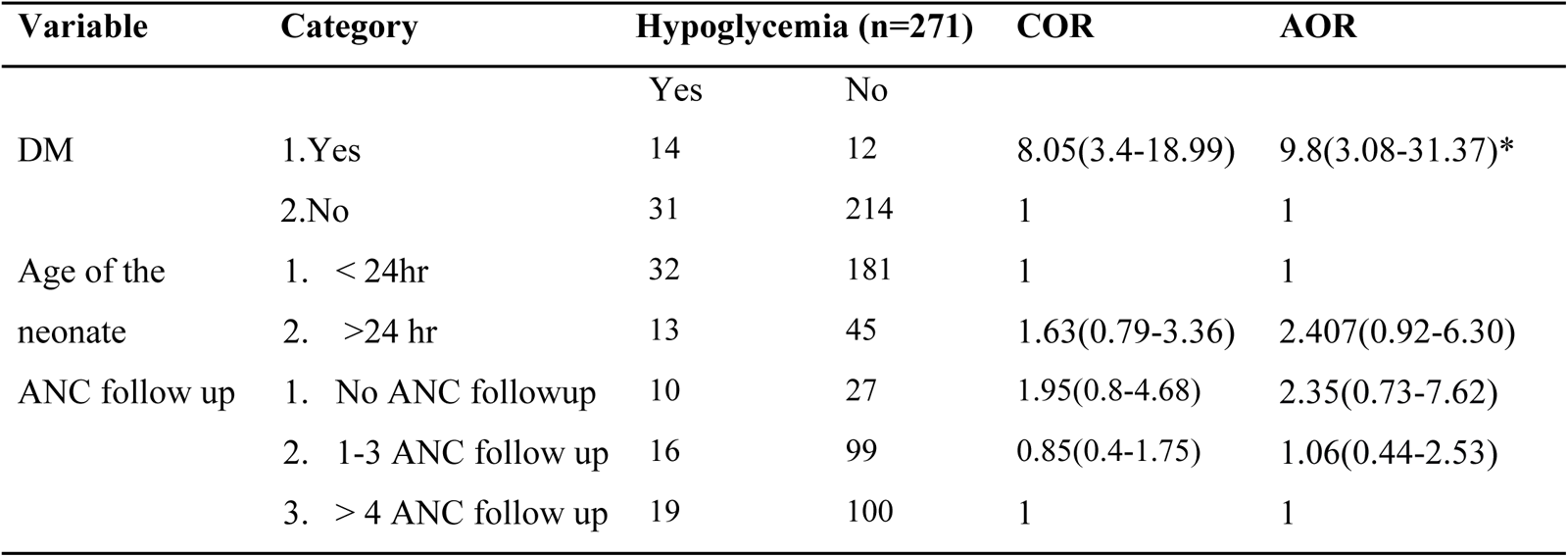
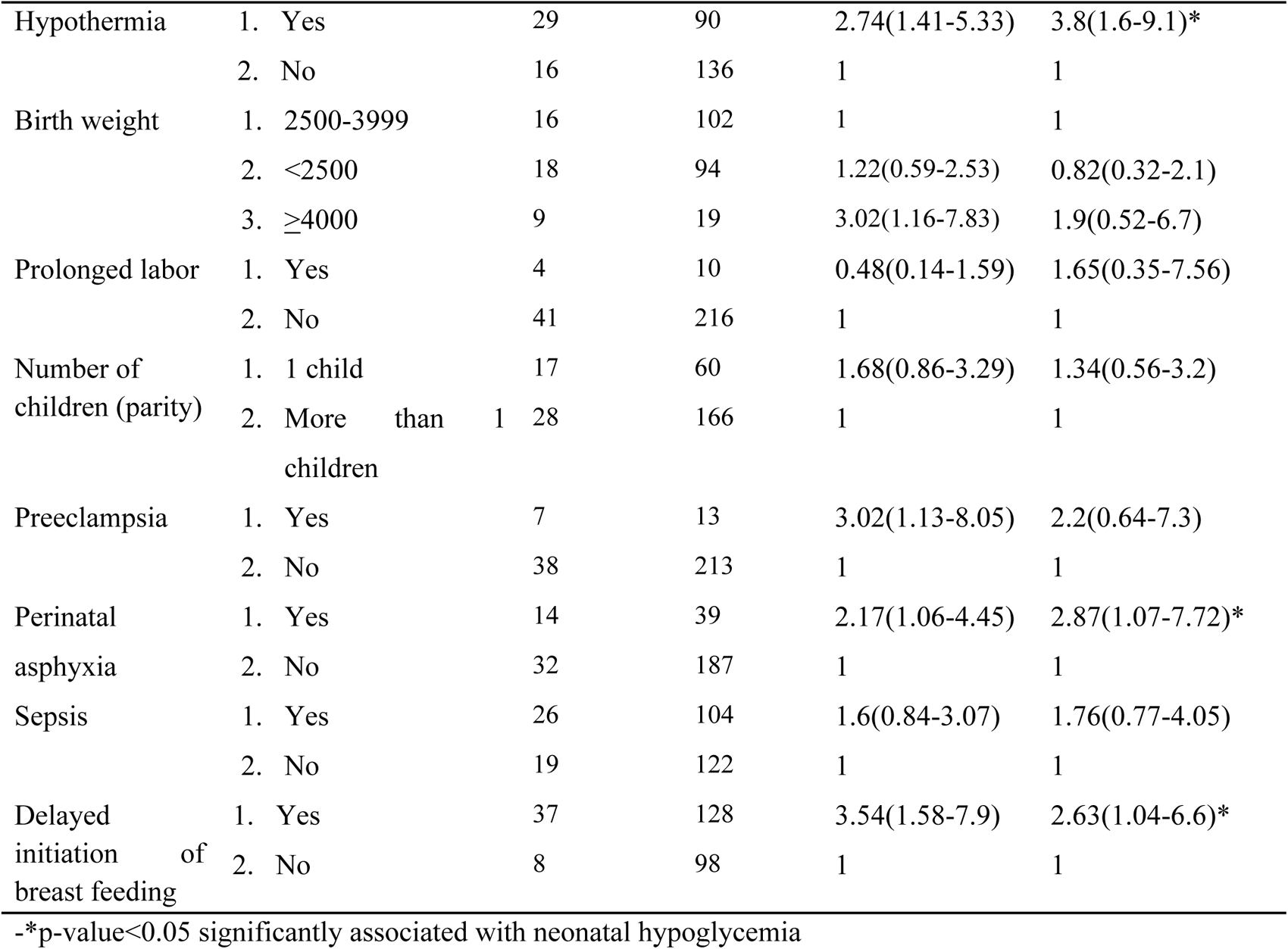
Factors associated with Hypoglycemia among neonates on admission to neonatal intensive care unit in Hawassa public Hospitals, Ethiopia 2023. (n=271)

| Variable | Category | Hypoglycemia (n=271) |  | COR | AOR |
| --- | --- | --- | --- | --- | --- |
|  |  | Yes | No |  |  |
| DM | 1.Yes | 14 | 12 | 8.05(3.4-18.99) | 9.8(3.08-31.37)* |
|  | 2.No | 31 | 214 | 1 | 1 |
| Age of the neonate | 1. < 24hr | 32 | 181 | 1 | 1 |
|  | 2. >24 hr | 13 | 45 | 1.63(0.79-3.36) | 2.407(0.92-6.30) |
| ANC follow up | 1. No ANC followup | 10 | 27 | 1.95(0.8-4.68) | 2.35(0.73-7.62) |
|  | 2. 1-3 ANC follow up | 16 | 99 | 0.85(0.4-1.75) | 1.06(0.44-2.53) |
|  | 3. > 4 ANC follow up | 19 | 100 | 1 | 1 |
| Hypothermia | 1. Yes | 29 | 90 | 2.74(1.41-5.33) | 3.8(1.6-9.1)* |
|  | 2. No | 16 | 136 | 1 | 1 |
| Birth weight | 1. 2500-3999 | 16 | 102 | 1 | 1 |
|  | 2. <2500 | 18 | 94 | 1.22(0.59-2.53) | 0.82(0.32-2.1) |
|  | 3. ≥4000 | 9 | 19 | 3.02(1.16-7.83) | 1.9(0.52-6.7) |
| Prolonged labor | 1. Yes | 4 | 10 | 0.48(0.14-1.59) | 1.65(0.35-7.56) |
|  | 2. No | 41 | 216 | 1 | 1 |
| Number of children (parity) | 1. 1 child | 17 | 60 | 1.68(0.86-3.29) | 1.34(0.56-3.2) |
|  | 2. More than 1 children | 28 | 166 | 1 | 1 |
| Preeclampsia | 1. Yes | 7 | 13 | 3.02(1.13-8.05) | 2.2(0.64-7.3) |
|  | 2. No | 38 | 213 | 1 | 1 |
| Perinatal asphyxia | 1. Yes | 14 | 39 | 2.17(1.06-4.45) | 2.87(1.07-7.72)* |
|  | 2. No | 32 | 187 | 1 | 1 |
| Sepsis | 1. Yes | 26 | 104 | 1.6(0.84-3.07) | 1.76(0.77-4.05) |
|  | 2. No | 19 | 122 | 1 | 1 |
| Delayed initiation of breast feeding | 1. Yes | 37 | 128 | 3.54(1.58-7.9) | 2.63(1.04-6.6)* |
|  | 2. No | 8 | 98 | 1 | 1 |

-\*p-value<0.05 significantly associated with neonatal hypoglycemia

## 5. Discussion

Hypoglycemia is one of a metabolic disturbance which occurs during the neonatal period. Screening at-risk infants and the management of low blood glucose levels in the first hours to days of life is a frequent issue in the care of newborn infants. If it is persistent and not detected early, it could lead to adverse neurological complications and poor prognosis. So this study was targeted to identify factors associated with neonatal hypoglycemia.

In this study, the prevalence of neonatal hypoglycemia at the admission point was 16.6% among neonates in the Neonatal intensive care unit, in Hawassa city. This study is in line with a study conducted in West Bengal and India which was 16.3% and 15.2% respectively (18, 19). On the other hand the finding was lower than the study carried out in Ethiopia St. Paul at 25% and Nigeria at 32.7 % (10, 12). This might be due to that these two studies used a higher cut-off point to diagnose neonatal hypoglycemia which is 47mg/dl.

In contrast, the magnitude of neonatal hypoglycemia was higher than the study conducted in USA(3.2%)(20), India(8.2%) (21), Nigeria (11%) (22), and Madagascar (3.1%) (23). The possible reasons may be, because our study includes infants of diabetic mothers, neonates born from hypertensive mothers, and newborns with severe congenital malformation which were excluded in the above study areas. Another study done in Uganda showed a 2.2% prevalence, which is much lower compared to the current study (6). The possible explanation for this may be that the study conducted in Uganda was community-based which is different from institutional based and large number of study participants involved in the study.

Regarding factors associated with neonatal Hypoglycemia, Diabetes mellitus (gestational Diabetes mellitus Pre-diabetes mellitus), perinatal asphyxia, delayed initiation of breastfeeding and the presence of Hypothermia were independently associated with it.

The current study finding revealed that neonates born from mothers with diabetes mellitus were nearly ten times more likely to have hypoglycemia compared with non-diabetic mothers. This study finding was supported by the study done in Ethiopia (Addis Ababa Black Loin Hospital and Harer Hiwot Fana Hospital), South Africa, Bangladesh, and China (11, 17, 24–26). This finding was can be justified by the fact that infants of diabetic mothers have hyper-insulinemia caused by high maternal glucose levels and after birth when maternal glucose is withdrawn the baby may develop hypoglycemia(27).

In contrast, the study carried out in St. Paul’s, Ethiopia, Indonesia and Israel, maternal DM was not found to be a contributing factor to neonatal hypoglycemia (12, 28, 29). The possible reason for maternal DM to be non-significant may be due to the intervention of early management of maternal DM. Another evidence for this variation may be due to absence or a small number of a mother with DM was found as a study participant.

Another factor that was found significantly associated with hypoglycemia was found to be perinatal asphyxia. Neonates born with perinatal asphyxia were nearly three times more likely to have hypoglycemia compared with non-asphyxia neonates. A similar result was also reported in Indonesia which showed that those who have perinatal asphyxia have a three times risk of having hypoglycemia(28). Evidence from china and Colombia suggests that hypoglycemia was significantly related to perinatal asphyxia (30)(Cristo Colmenares et al., 2021). This may be due to the reason in perinatal asphyxia, increasing metabolic and energy demands are achieved transiently by increasing glucose production through glycogenolysis. Due to hypoxia, the infant switches to anaerobic metabolism, resulting in rapid exhaustion of glycogen stores, hypoglycemia, and lactic acedemia(31).

Hypothermia was found to be one of the significant risk factors associated with hypoglycemia in this study. Neonates with hypothermia were four times more likely to have hypoglycemia than non-hypothermic. Various evidence show that hypothermia is one of the major risk factor for hypoglycemia (11, 22, 24, 26). In a cross-sectional study in Ethiopia, Hiwot Fana Hospital and St. Paulo’s Hospital the odds of developing risk of hypoglycemia in hypothermic neonates were three and two respectively (11, 12). After birth maintaining newborn body temperature is very important. When the baby gets cold the newborn uses up more glycogen to keep warm. Then the newborn must utilize glucose stores to keep warm, then the blood sugar drops and they become hypothermic along with hypoglycemic(32).

Delayed initiation of breastfeeding is a major contributing factor to neonatal hypoglycemia. Newborns that started initial breastfeeding after one hour had a three times higher chance of developing neonatal hypoglycemia compared with those who started within one hour. Similarly, research conducted in China, Uganda and Ethiopia Harer, late initiation of breastfeeding has shown that three times higher risk of developing hypoglycemia (6, 11, 24). This similarity can be due to the reason that neonates with delayed initiation of breastfeeding lacks the advantage early initiation of breastfeeding i.e. colostrum increases the blood glucose level within one hour and continuing breast feeding maintain infants at eu-glycemic state(33).

## 6. Strengths and Limitations of the Study

This study used primary data for blood glucose measurement, so it reduces information bias. The limitation of this study is an institution based study it does not address the general community and the study doesn’t incorporate laboratorial investigations like RFT, LFT, Bilirubin count etc.

## 7. Conclusion

In this study, the magnitude of hypoglycemia was 16.6% which is high. The result of this study proved that maternal DM, delayed initiation feeding, hypothermia and perinatal asphyxia were associated with the increasing risk of neonatal hypoglycemia. Therefore, early initiation of feeding is an important factor to prevent hypoglycemia for all newborns. On the other hand, preventing, early detection and management of hypoglycemia should be done in neonates with hypothermia and perinatal asphyxia.

## Data Availability

All relevant data are within the manuscript and its Supporting Information files.

## Acknowledgment

We the authors acknowledged study participants, data collectors, supervisors, Hawassa City Administration Health Bureau, and public hospitals for feeding the necessary information and their unwavering efforts in the research process. Moreover, we appreciate Hawassa University to pursue this chance to conduct this research and Dilla University for covering the data collection cost.

## Reference

1. stanford m. Hypoglycemia in a Newborn Baby 2023 [cited 2023 march, 2]. Available from: https://www.stanfordchildrens.org/en/topic/default?id=hypoglycemia-in-the-newborn-90-P01961.

2. Dixon KC, Ferris RL, Marikar D, Chong M, Mittal A, Manikam L, et al. Definition and monitoring of neonatal hypoglycaemia: a nationwide survey of NHS England Neonatal Units. Archives of Disease in Childhood-Fetal and Neonatal Edition. 2017;102(1):F92–F3.

3. Harris DL, Weston PJ, Battin MR, Harding JE. A survey of the management of neonatal hypoglycaemia within the Australian and New Zealand Neonatal Network. Journal of Paediatrics and Child Health. 2014;50(10):E55–E62.

4. EMOH. neonatal intensive care unit management protocol 2021.

5. Stanescu A, Stoicescu S. Neonatal hypoglycemia screening in newborns from diabetic mothers-Arguments and controversies. Journal of medicine and life. 2014;7(Spec Iss 3):51.

6. Mukunya D, Odongkara B, Piloya T, Nankabirwa V, Achora V, Batte C, et al. Prevalence and factors associated with neonatal hypoglycemia in Northern Uganda: a community-based cross-sectional study. Tropical Medicine and Health. 2020;48(1):1–8.

7. Hay WW, Raju TN, Higgins RD, Kalhan SC, Devaskar SU. Knowledge gaps and research needs for understanding and treating neonatal hypoglycemia: workshop report from Eunice Kennedy Shriver National Institute of Child Health and Human Development. The Journal of pediatrics. 2009;155(5):612–7.

8. Harding JE, Harris DL, Hegarty JE, Alsweiler JM, McKinlay CJ. An emerging evidence base for the management of neonatal hypoglycaemia. Early human development. 2017;104:51–6.

9. Shah R, Harding J, Brown J, McKinlay C. Neonatal Glycaemia and Neurodevelopmental Outcomes: A Systematic Review and Meta-Analysis. Neonatology. 2019;115(2):116–26.

10. Dedeke I, Okeniyi J, Owa J, Oyedeji G. Point-of-admission neonatal hypoglycaemia in a Nigerian tertiary hospital: incidence, risk factors and outcome. Nigerian Journal of Paediatrics. 2011;38(2):90–4.

11. Sertsu A, Nigussie K, Eyeberu A, Tibebu A, Negash A, Getachew T, et al. Determinants of neonatal hypoglycemia among neonates admitted at Hiwot Fana Comprehensive Specialized University Hospital, Eastern Ethiopia: A retrospective cross-sectional study. SAGE Open Medicine. 2022;10:20503121221141801.

12. Fantahun BY, Nurussen I. Prevalence and Risk factors of Hypoglycaemia in Neonates at St. Paul’s Hospital Millennium Medical College Neonatal Intensive Care Unit, Ethiopia: A Cross Sectional Study. 2020.

13. Amponsah G, Hagan O, Okai E. Neonatal Hypoglycaemia at Cape Coast Teaching Hospital. Journal of the West African College of Surgeons. 2015;5(2):100.

14. HUCSHHR hucshhr. Number of serving population and staff profile. 2023.

15. AGHHR aghhr. Number of serving population and staff profile. 2023.

16. TPHHR TpH. Number of serving population and staff profile. 2023.

17. Fikirte Kasaye RM, Kerebih Abere. Prevalence and predictors of hypoglycemia among neonates on admission to NICU in Addis Ababa public Hospitals, Addis Ababa, Ethiopia, 2021. Addis abeba university repository 2021.

18. Meyur R, Sadhu A, Bhakta A, Bandyopadhyay M, Kundu B, Bhaumik S, et al. INCIDENCE & CAUSES OF NEONATAL HYPOGLYCEMIA AFTER CESAREAN SECTION IN A RURAL SETUP OF WEST BENGAL. Journal of Evolution of medical and Dental Sciences. 2014;3:1191–4.

19. Singh YP, Devi TR, Gangte D, Devi TI, Singh NN, Singh MA. Hypoglycemia in newborn in Manipur. Journal of Medical Society. 2014;28(2):108–11.

20. Hosagasi NH, Aydin M, Zenciroglu A, Ustun N, Beken S. Incidence of hypoglycemia in newborns at risk and an audit of the 2011 American academy of pediatrics guideline for hypoglycemia. Pediatrics & Neonatology. 2018;59(4):368–74.

21. Babu M, D’Souza J, Susheela C. Study of incidence, clinical profile and risk factors of neonatal hypoglycemia in a tertiary care hospital. Int J Pediatr Res. 2016;3(10):753.

22. Ochoga MO, Aondoaseer M, Abah RO, Ogbu O, Ejeliogu EU, Tolough GI. Prevalence of Hypoglycaemia in Newborn at Benue State University Teaching Hospital, Makurdi, Benue State, Nigeria. Open Journal of Pediatrics. 2018;8(2):189–98.

23. Sambany E, Pussard E, Rajaonarivo C, Raobijaona H, Barennes H. Childhood dysglycemia: prevalence and outcome in a referral hospital. PloS one. 2013;8(5):e65193.

24. Zhao T, Liu Q, Zhou M, Dai W, Xu Y, Kuang L, et al. Identifying risk effectors involved in neonatal hypoglycemia occurrence. Bioscience Reports. 2020;40(3).

25. Magadla Y, Velaphi S, Moosa F. Incidence of hypoglycaemia in late preterm and term infants born to women with diabetes mellitus. South African Journal of Child Health. 2019;13(2):78–83.

26. Hassan M, Pervez A, Biswas R, Debnath S, Syfullah K. Incidence and Risk Factors of Neonatal Hypoglycemia During the First 48 Hours of Life in a Tertiary Level Hospital. Faridpur Medical College Journal. 2020;15:12–5.

27. Bamehrez M. Hypoglycemia and associated comorbidities among newborns of mothers with diabetes in an academic tertiary care center. Frontiers in Pediatrics. 2023;11.

28. Yunarto Y, Sarosa GI. Risk factors of neonatal hypoglycemia. Paediatrica Indonesiana. 2019;59(5):252–6.

29. Bromiker R, Perry A, Kasirer Y, Einav S, Klinger G, Levy-Khademi F. Early neonatal hypoglycemia: incidence of and risk factors. A cohort study using universal point of care screening. The Journal of Maternal-Fetal & Neonatal Medicine. 2019;32(5):786–92.

30. Zhou W, Yu J, Wu Y, Zhang H. Hypoglycemia incidence and risk factors assessment in hospitalized neonates. The Journal of Maternal-Fetal & Neonatal Medicine. 2015;28(4):422–5.

31. Chandran S, Samuel Rajadurai V, Alim Abdul Haium A, Hussain K. Current perspectives on neonatal hypoglycemia, its management, and cerebral injury risk. Research and reports in Neonatology. 2015:17–30.

32. Senadhi V, Dutta S. Recurrent Hypothermia and Hypoglycemia as the Initial Presentation of Hepatitis C: 724. Official journal of the American College of Gastroenterology | ACG. 2010;105:S262.

33. Cordero L, Stenger MR, Landon MB, Nankervis CA. Early feeding, hypoglycemia and breastfeeding initiation in infants born to women with pregestational diabetes mellitus. J Neonatal Perinatal Med. 2018;11(4):357–64.

